# The Association Between Plasma Omega-3 Fatty Acids and Suicidal Ideation/Self-harm in the United Kingdom Biobank

**DOI:** 10.1101/2025.04.11.25325688

**Authors:** W Grant Franco, Nathan Tintle, Jason Westra, Evan L O’Keefe, James H O’Keefe, William S Harris

## Abstract

**Background:** Self-harm is a significant public health concern, with increasing prevalence globally. Omega-3 fatty acids (FAs), particularly eicosapentaenoic acid (EPA) and docosahexaenoic acid (DHA), are known for their health benefits, including potential mental health improvements. This study explores the association between omega-3 FAs and self-harm behaviors using data from the UK Biobank (UKBB).

**Objectives:** To investigate the relationship between plasma levels of omega-3 FAs and various measures of self-harm, including passive suicidal ideation and deliberate self-harm, within a large cohort study.

**Methods:** This observational study analyzed data from a random subset of 258,012 participants with plasma omega-3 FA levels, covariate data, and self-harm records. Omega-3 levels were measured using Nuclear Magnetic Resonance (NMR) and are expressed as a percent of total FAs. Self-harm outcomes were assessed through self-reported questionnaires and medical records. Covariates included demographic, health, and lifestyle factors. Statistical analyses involved logistic regression and Cox proportional hazards models, adjusting for relevant covariates. Adjusted odds ratios (aORs) are presented and 95% confidence intervals.

**Results:** Higher levels of DHA, ALA+EPA+DPA and total omega-3 were generally inversely associated with passive suicidal ideation, history of self-harm, and future self-harm, with DHA showing the strongest associations. Participants in the highest quintile of DHA had a 13% lower risk of passive suicidal ideation (aOR = 0.87; 95% CI 0.82, 0.92), and lower odds of history of self-harm compared to those in the lowest quintile (aOR = 0.67; 95% CI 0.55, 0.83). These associations were generally stronger for medical record-based outcomes than for self-reported data.

**Conclusions:** This study provides evidence that higher omega-3 FA levels, particularly DHA, are associated with reduced risks of self-harm and suicidal ideation. These findings suggest that omega-3 FAs may play a protective role in mental health, highlighting the potential of dietary interventions to mitigate self-harm behaviors.

## INTRODUCTION

The prevalence of self-harm, a significant public health concern globally, has increased over recent years. A recent report from England showed a two-fold increase in reported self-harm from 2.4% prevalence to 6.4% between 2000 and 2012 with a particularly notable increase among females aged 16-24 years, where prevalence jumped from 6.5% to 19.7% (1). Deliberate self-harm, which includes non-suicidal self-injury (NSSI) involves behaviors such as cutting, burning, or self-poisoning and is often a manifestation of psychological distress (2–4). Studies have reported varying rates of self-harm across different populations, with lifetime prevalence of NSSI of 22% (5–8). These findings underscore the urgency of addressing self-harm and its underlying causes to mitigate its impact on individuals and society.

Omega-3 fatty acids (FAs), in particular eicosapentaenoic (EPA) and docosahexaenoic acids (DHA), are bioactive constituents of certain fish and seafoods and have well-documented health benefits. These include protective effects against cardiovascular disease (CVD) and cognitive decline (9). Recent evidence suggests that omega-3 FAs may also offer significant mental health benefits, potentially reducing the risk of depression, and anxiety, and possibly influencing self-harm behaviors (10). The etiology of these benefits is believed to be linked to the anti-inflammatory properties of these FAs and to their crucial role in brain function (11). Specifically, EPA and DHA are integral to maintaining neuronal cell membrane fluidity and function, which in turn supports neurotransmission and may mitigate mood disorders (12).

Supporting evidence for the impact of omega-3 FAs on mental health is growing, with studies linking higher levels of these nutrients to reduced symptoms of depression and anxiety (13–15). For instance, the Framingham Heart Study found that higher omega-3 index levels were associated with significantly lower risks of major depressive disorder (MDD) and other psychiatric conditions (16). Moreover, omega-3 supplementation has shown promise in reducing the severity of symptoms in individuals with clinical depression, suggesting a potential therapeutic role for these FAs in managing mental health conditions (13–15). A meta-analysis of RCTs showed a possible use of omega-3 supplementation as adjuvant therapy in MDD (17).

There is evidence that omega-3 FA supplementation may provide benefit for patients with a history of self-injury and suicidal behavior (9,18–26), however the evidence is unclear with other studies showing a lack of predictive power for self-harm or suicide (18,27,28). Together, these findings suggest that omega-3 FAs may be beneficial across a range of mental health issues, including self-harm, but these relationships are still not widely accepted (29,30). Thus, in this study our goal is to determine the relationships between plasma levels of omega-3 FAs and measures of prevalent self-harm in the United Kingdom Biobank (UKBB). Our hypothesis was that higher omega-3 FA levels would be associated with lower risk for measures of self-harm.

## METHODS

### Sample

This observational study was conducted within the UKBB, a prospective, population-based cohort of 502,366 individuals, 40-69 years of age, recruited in the UK between April 2007 and December 2010. UKBB has ethical approval (Ref. 11/NW/0382) from the Northwest Multi-Centre Research Ethics Committee as a Research Tissue Bank. The University of South Dakota Institutional Review Board approved the use of these de-identified, publicly available data for research purposes (IRB-21-147). Within the cohort, a random subset of 258,012 participants had data on plasma omega-3 FA levels (exposures) and medical-record based information on self-harm or self-reported self-harm (outcomes) as well as complete data on relevant covariates and serves as our primary analytic sample. In secondary analyses, we also examined relationships in a subset of 80,745 individuals who answered more detailed questions on lifetime and 12-month outcome variables. The median follow-up time was 13.7 years (max=15.8 years).

### Variables of Interest

#### Exposures

The plasma omega-3 FA levels were measured by Nuclear Magnetic Resonance (NMR) (Nightingale Health Plc, Helsinki, Finland) and expressed as percent of total FAs. The primary measures reported were total omega-3% and DHA%. We derived a third variable (alpha linolenic acid [ALA] + EPA + docosapentaenoic acid [DPA]%), by subtracting DHA% from total omega-3%.

### Outcomes

We looked at 3 sets of outcomes (9 total variables) related to suicidal ideation, self-harm and tendencies to these behaviors. The first set of 3 variables were self-reported answers to questions about life being worth living (passive suicidal ideation) and contemplations of self-harm, both lifetime and in the previous 12-months. The second set of 4 variables were related to history of actual self-harm: self-reported history (ever; past 12-months), and as noted in the medical-record (ever; past 12-months). The third set of 2 variables were derived from participant medical records and involved future events: (a) first time self-harm (i.e., after baseline, among people with no-history of self-harm), and (b) any self-harm (after baseline, among all people, regardless of history of self-harm). Loss to follow-up was censored at last data measurement.

The exact verbiage of the questions, and ICD-10 codes used from the medical record are provided in Supplemental Table 1a.

### Covariates

We considered the following covariates: self-reported biological sex (male/female), age (years), body mass index (BMI), marital status (married/partnered or not), employment status (disabled, employed, retired, unemployed, other), self-reported race/ethnicity (White, other), self-reported exercise (quartiles of moderate-to-vigorous exercise; as minutes/week), Alcohol consumption (self-reported days/week), history of depression, history of anxiety disorder, smoking (pack years), psychotropic medication use (yes/no), comorbid disorders (count) and the Townsend Deprivation Index. Additional details on variable definitions are provided in Supplemental Table 1b.

### Statistical Analyses

Sample characteristics are summarized using standard statistical methods, including means, standard deviations (SD), and %s. A mix of logistic regression (cross-sectional – lifetime and 12-month) models and Cox proportional hazards models (future outcomes) were used.

Models were fit for continuous versions of the three omega-3 FA exposures per interquintile range (IQ_5_R, defined as the difference between the 10^th^ and 90^th^ percentiles). Separately, each FA was also modeled categorically with separate effect estimates for each quintile. Three PUFA exposures (DHA%, ALA+EPA+DPA% and total omega-3%) and 9 outcomes were considered, for a total of 27 models, all adjusted for the covariates described above. Cubic splines using 3 knots (at 25^th^, 50^th^ and 75^th^ percentiles), and after dropping the top/bottom 2.5% of the data were fit in all cases, and then compared to continuous/linear models using Wald tests. Statistical significance was set to 0.05 and 95% confidence intervals (CI) are provided for most comparisons. Statistical significance for tests of non-linearity was set to 0.01 to account for multiple testing. R (www.r-project.org) was used for all analyses.

## Results

The UKBB sample is slightly more female than male, primarily white and with a mean age of slightly over 56 years. Other characteristics of the sample are shown in **Table 1**. Overall, there were only minor differences between the sample with medical-records data and the smaller set of individuals who also had self-reported mental health/ behavior data. However, the mean (SD) DHA % was slightly lower in the medical record sample (1.99% [0.66]) vs the self-report sample (2.06% [0.66]), with a similar pattern for ALA+EPA+DPA (2.39 [0.99] vs. 2.43 [0.99]) and total omega-3 (4.39 [1.55] vs. 4.50 [1.56]).

**Table 1.** Sample Descriptives.

| Variable | Category | Medical record-based analyses<br>N=260,784 | Self-reported<br>mental health<br>and behavior<br>analyses<br>N=81,024 |
| --- | --- | --- | --- |
|  |  | Mean (SD) or % (X) |  |
| Sex | Female | 53.0 (136760) | 54.9 (44303) |
| Age |  | 56.7 (8.1) | 56.1 (7.7) |
| Self-reported<br>Race/Ethnicity | White | 95.5 (246439) | 97.4 (78615) |
| BMI |  | 27.5 (4.8) | 26.8 (4.6) |
| Marital Status | Married | 72.8 (187876) | 74.9 (60494) |
| Employment | Disabled | 3.4 (8705) | 1.5 (1225) |
|  | Employed | 59.1 (152368) | 65.1 (52579) |
|  | Other | 1.5 (3782) | 1.4 (1102) |
|  | Retired | 34.5 (89050) | 30.8 (24893) |
|  | Unemployed | 1.6 (4107) | 1.2 (946) |
| Townsend Deprivation<br>Index |  | -1.4 (3) | -1.8 (2.8) |
| Alcohol Consumption | Daily | 20.5 (52792) | 23.3 (18821) |
|  | Rarely | 29.8 (76879) | 25.1 (20242) |
|  | One to two times/week | 26.3 (67735) | 25.2 (20349) |
|  | Three to four times/week | 23.5 (60606) | 26.4 (21333) |
| Smoking Pack Years |  | 10.6 (15.6) | 8.9 (13.6) |
| Exercise | Highest > 3,393 METs/wk | 23.1 (59544) | 25.9 (20884) |
|  | High >1,618 METs/wk | 23.3 (59994) | 21.3 (17216) |
|  | Low >690 METs/wk | 23 (59357) | 26 (20959) |
|  | Lowest <690 METs/wk | 22.5 (57975) | 22.6 (18224) |
|  | Missing | 8.2 (21142) | 4.3 (3462) |
| History of anxiety | Yes | 1.9 (4869) | 1.6 (1331) |
| History of Depression | Yes | 34.1 (88036) | 32.9 (26582) |
| Comorbidities | 0 | 46.1 (118830) | 51.2 (41378) |
|  | 1 | 33.2 (85758) | 32.6 (26325) |
|  | 2 | 14 (36009) | 11.9 (9595) |
|  | 3+ | 6.7 (17415) | 4.3 (3447) |
| Psychotropic medication<br>use | Yes | 2.8 (7243) | 2.1 (1664) |

### Past passive suicidal ideation and contemplating self-harm

The self-reported lifetime prevalence of having perceived one’s life as not worth living (passive suicidal ideation) was high (30.5%) (**Table 2**). DHA, total omega-3 and ALA+EPA+DPA levels all showed strong inverse associations with this metric. DHA showed the strongest evidence of inverse association with passive suicidal ideation with a 13% reduction in risk when comparing the highest to the lowest quintiles of DHA in the multivariable adjusted model. DHA also showed significant evidence of non-linearity (p=0.003). Subsequent cubic spline analysis of the DHA vs prevalent passive suicidal ideation revealed an increase in risk below a DHA level of about 1.7% (**Figure 1**). Similar associations with this outcome were observed for total omega-3 PUFA levels (although the magnitude of associations were slightly less), while inverse associations with ALA+EPA+DPA were even smaller, though still statistically significant (**Table 2**), and did not have statistically significant evidence of non-linearity (p>0.01). Lifetime and past 12-month associations with omega-3 levels and contemplations of self-harm also showed generally inverse relationships with a mix of statistically significant and non-significant findings, and risk reductions between 0 and 14% when comparing Q2-Q5 to Q1. None of these comparisons showed statistically significant evidence of non-linearity (p>0.01 in all cases).

**Figure 1.**
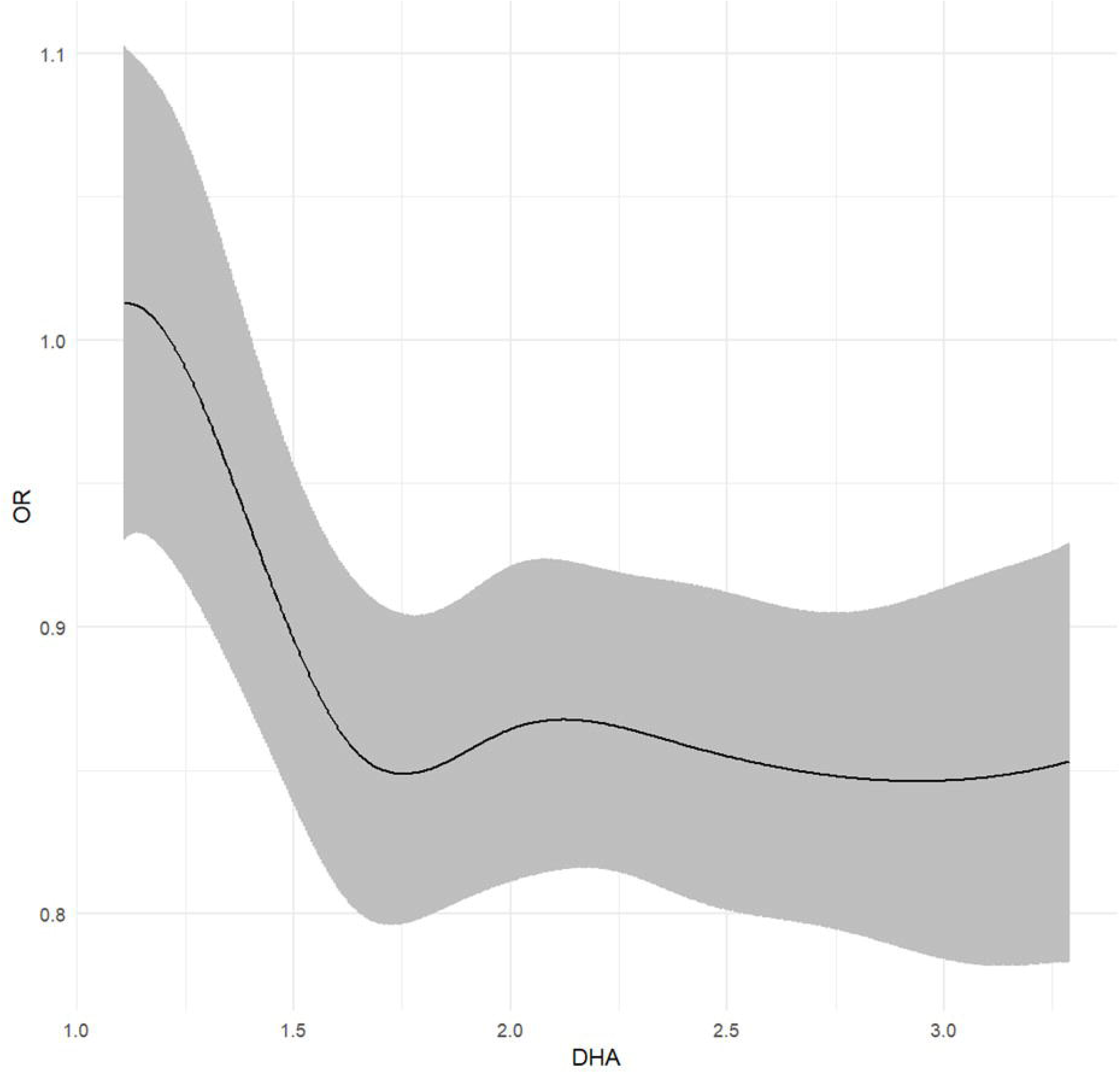
Cubic spline for DHA (%) vs. odds of passive suicidal ideation

**Table 2.** Associations of omega-3 levels with lifetime self-reported contemplations of self-harm and passive suicidal ideation^1^.

|  |  |  | Ever passive suicidal ideation<br>(N=81,024) |  | Ever contemplate Self-harm<br>(N=81,024) |  | Contemplate Self-Harm<br>(in the last year)<br>(N=81,024) |  |
| --- | --- | --- | --- | --- | --- | --- | --- | --- |
|  |  | n | % (x) | aOR (95% CI) | % (x) | aOR (95% CI) | % (x) | aOR (95% CI) |
| **DHA** | per IQ<sub>5</sub>R | 80745 | 30.5%<br>(24609) | 0.93 (0.89,0.97)*** | 14.3%<br>(11553) | 0.96<br>(0.91,1.02) | 2.8%<br>(2273) | 0.93<br>(0.82,1.04) |
|  | Q1<br>(<1.46%) | 13390 | 33.5%<br>(4488) | NA | 16.2%<br>(2164) | NA | 3.6%<br>(476) | NA |
|  | Q2 (1.46-1.78%) | 14981 | 30.9%<br>(4623) | 0.89 (0.84,0.94)*** | 14.6%<br>(2192) | 0.91<br>(0.85,0.98)* | 3%<br>(454) | 0.94<br>(0.82,1.08) |
|  | Q3 (1.78-2.07%) | 16217 | 30.3%<br>(4912) | 0.87 (0.83,0.92)*** | 14.0%<br>(2266) | 0.89<br>(0.83,0.96)** | 2.9%<br>(477) | 0.98<br>(0.86,1.13) |
|  | Q4 (2.07-2.47%) | 17628 | 29.9%<br>(5263) | 0.89 (0.84,0.94)*** | 14.1%<br>(2493) | 0.96<br>(0.90,1.03) | 2.6%<br>(460) | 0.95<br>(0.82,1.09) |
|  | Q5<br>(>2.47%) | 18529 | 28.7%<br>(5323) | 0.87 (0.82,0.92)*** | 13.2%<br>(2438) | 0.94<br>(0.88,1.01) | 2.2%<br>(406) | 0.89<br>(0.77,1.04) |
| **Total**<br>**Omega-3** | per IQ<sub>5</sub>R | 80745 | 30.5%<br>(24609) | 0.94 (0.91,0.98)** | 14.3%<br>(11553) | 0.95<br>(0.90,1.00)* | 2.8%<br>(2273) | 0.90<br>(0.81,1.01) |
|  | Q1<br>(<3.16%) | 14405 | 33.8%<br>(4872) | NA | 16.9%<br>(2438) | NA | 3.6%<br>(525) | NA |
|  | Q2 (3.16-3.85%) | 15308 | 31.5%<br>(4826) | 0.95 (0.91,1.01) | 15.1%<br>(2315) | 0.95<br>(0.89,1.01) | 3.2%<br>(492) | 1.00<br>(0.87,1.14) |
|  | Q3 (3.85-4.52%) | 16192 | 30.3%<br>(4904) | 0.94 (0.89,0.99)* | 14.0%<br>(2274) | 0.92<br>(0.86,0.98)* | 2.9%<br>(465) | 0.96 (0.84,1.1) |
|  | Q4 (4.52-5.46%) | 17106 | 29.2%<br>(4999) | 0.91(0.87,0.96)*** | 13.2%<br>(2265) | 0.90<br>(0.84,0.97)** | 2.4%<br>(412) | 0.89<br>(0.77,1.02) |
|  | Q5 | 17734 | 28.2% | 0.92 (0.87,0.97)** | 12.7% | 0.95 | 2.1% | 0.90 |
|  | (>5.46%) |  | (5008) |  | (2261) | (0.88,1.01) | (379) | (0.78,1.04) |
| **ALA+EPA+DPA** | per IQ<sub>5</sub>R | 80745 | 30.5%<br>(24609) | 0.95 (0.92,1.00)* | 14.3%<br>(11553) | 0.94<br>(0.89,0.99)* | 2.8%<br>(2273) | 0.90 (0.8,1.01) |
|  | Q1<br>(<1.57%) | 15364 | 33.5%<br>(5154) | NA | 17.1%<br>(2632) | NA | 3.8%<br>(581) | NA |
|  | Q2 (1.57-2.08%) | 15774 | 31.5%<br>(4963) | 0.99 (0.94,1.04) | 14.9%<br>(2347) | 0.93<br>(0.87,1.00)* | 2.9%<br>(455) | 0.86<br>(0.75,0.98)* |
|  | Q3 (2.08-2.54%) | 15980 | 30.2%<br>(4826) | 0.96 (0.91,1.02) | 13.8%<br>(2212) | 0.90<br>(0.84,0.97)** | 2.7%<br>(439) | 0.88<br>(0.77,1.01) |
|  | Q4 (2.54-3.14%) | 16548 | 29.2%<br>(4839) | 0.96 (0.91,1.01) | 13.4%<br>(2224) | 0.93<br>(0.87,0.99)* | 2.6%<br>(422) | 0.89<br>(0.78,1.02) |
|  | Q5<br>(>3.14%) | 17079 | 28.3%<br>(4827) | 0.95 (0.90,1.00)* | 12.5%<br>(2138) | 0.91<br>(0.85,0.98)** | 2.2%<br>(376) | 0.86<br>(0.75,0.99)* |

<sup>†</sup>Models were adjusted for all variables in Table 1.

### Lifetime self-reported/medical record inferred self-harm

**Figure 2** and **Supplemental Table 2a** provide odds ratios for the relationship between history of self-harm (self-reported and medical record inferred) and omega-3 PUFA levels.

**Figure 2.**
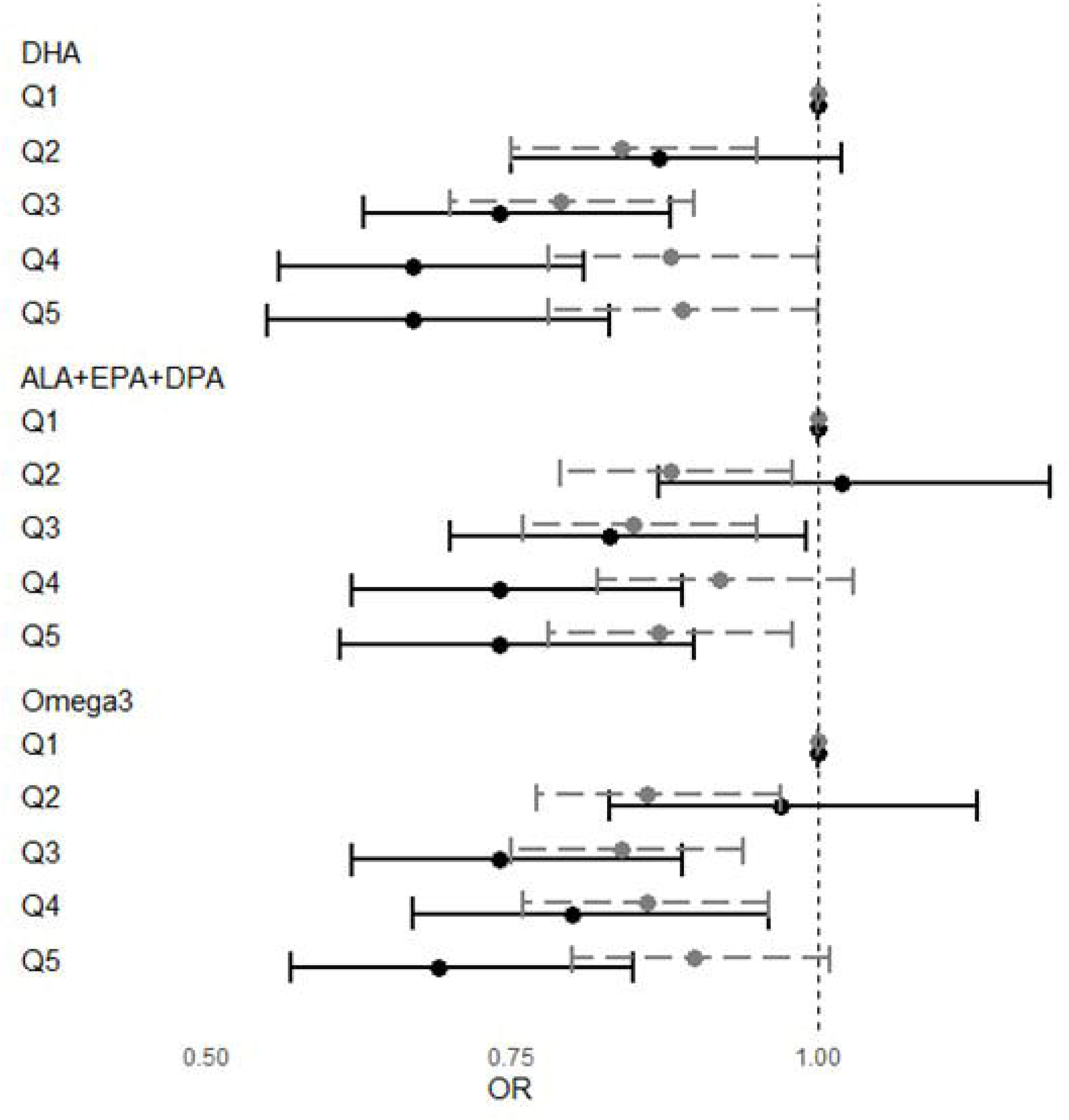
Associations between omega-3 PUFA levels and history of self-harm (self-report [gray] and medical record based [black])

Overall, associations were statistically significant in most cases, with the strongest associations for history of self-harm as inferred from the medical record (e.g., DHA per IQ_5_R OR 0.71 [95% CI: 0.60, 0.83]). Associations were less strong, and periodically not statistically significant, for self-report history of self-harm. Twelve-month history of self-harm had generally low prevalence (Supplemental Table 2b) but did show trends towards inverse associations with omega-3 levels, though many of the aHRs were not statistically significant. Tests of nonlinearity were not statistically significant (p>0.01).x

### Future self-harm

Associations between first time and any incident self-harm were also inversely associated with omega-3 FA levels in all cases. While overall prevalence was low (0.3-0.4% of the sample), comparisons between the highest and lowest quintiles of omega-3 levels showed HR between 0.71 and 0.83 showing a strong inverse relationship which was statistically significant (p<0.05) in most cases (**Table 3**). As with other outcomes, association showed no evidence of non-linearity using cubic splines (p>0.01 in all cases).

**Table 3.**
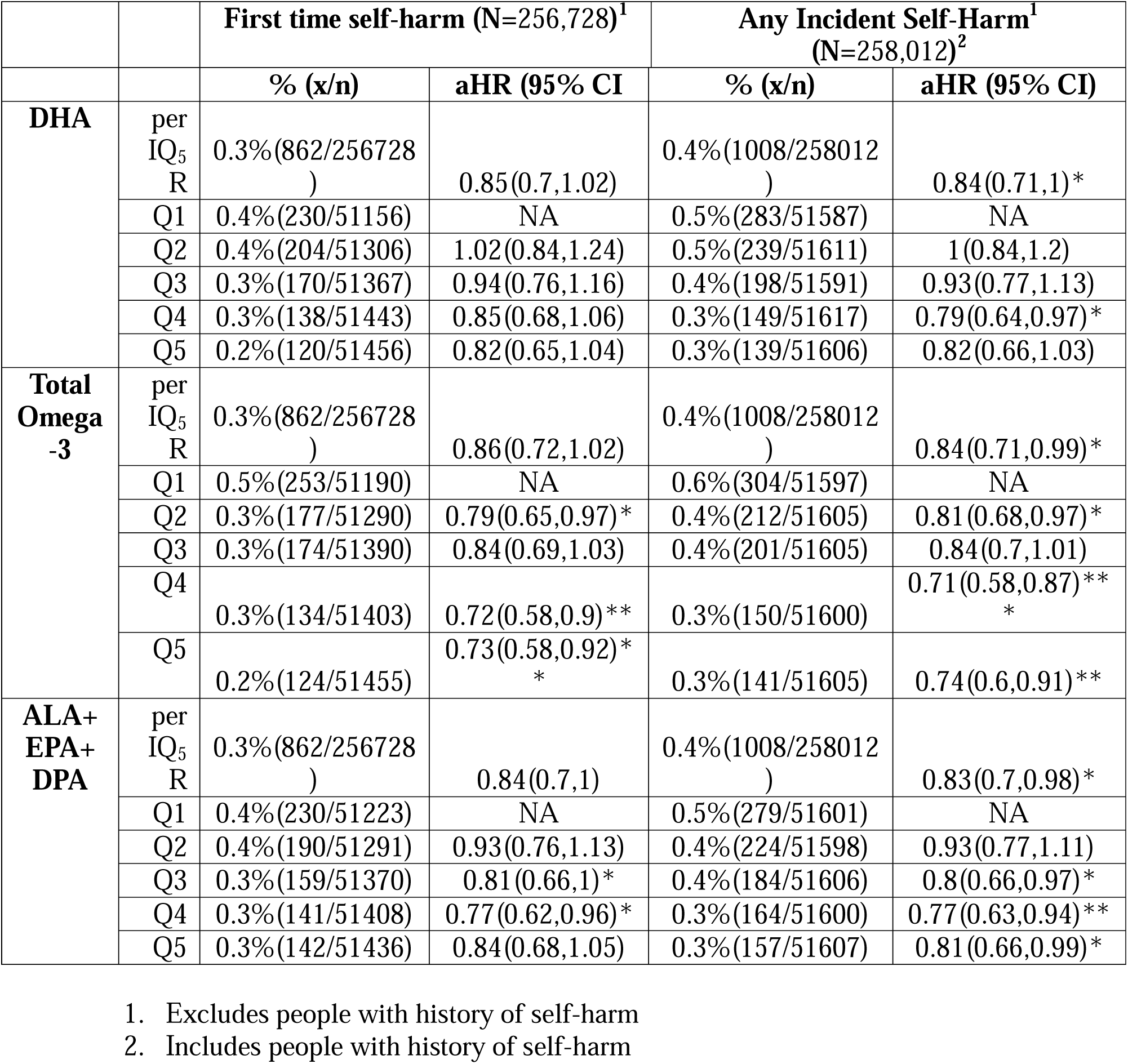
Associations between first time and any incident self-harm and omega-3 PUFA levels.

## Discussion

This study examined the association between omega-3 FA levels and self-harm/suicidal ideation using data from the UKBB cohort. We found an inverse relationship between higher omega-3 FA levels, particularly DHA, and both past and future self-harm and/or suicidal ideation. These results underscore the potential importance of omega-3 FA in mental health interventions, especially in mitigating risks associated with self-harm and suicidal behaviors.

We used 3 different outcomes in this study – passive suicidal ideation which was assessed through self-reporting “if life was worth living,” history of self-harm both assessed through self-reporting and medical records, and incident self-harm (both in first time and in all participants) from the medical records.

### Passive suicidal ideation

Our analysis showed that higher levels of DHA and total omega-3 were associated with a reduced risk of passive suicidal ideation. Participants in the highest quintile of DHA levels had an 11% lower risk of reporting that life was not worth living compared to those in the lowest quintile. This supports existing literature suggesting that omega-3 FA play a role in mood regulation and mental health (31–34).

### Self-Harm Reduction

We also found that higher DHA levels were inversely associated with both lifetime and past 12-month self-reported self-harm, though the associations with 12-month self-harm were less statistically significant. Medical record-based self-harm data reinforced these findings, showing a stronger inverse relationship. These results align with previous studies indicating that omega-3 supplementation can reduce symptoms of depression and potentially lower self-harm incidents (13–15,19).

### Future Self-Harm

The analysis of future self-harm (both first time events and any incident event regardless of a history of self-harm) revealed a strong inverse relationship with omega-3 levels. Hazard ratios (HR) for comparisons between the highest and lowest quintiles of omega-3 levels indicated a significant protective effect, suggesting that maintaining higher omega-3 levels could be a preventive strategy against future self-harm.

The mechanisms underlying the observed associations likely involve several factors. Omega-3, particularly DHA, is crucial for maintaining neuronal cell membrane fluidity, which is essential for proper neurotransmission and brain function (35,36). Because neuroinflammation has been linked to depression and other mental health disorders, Omega-3’s anti-inflammatory properties may play a role in improving brain health (36). Similarly, these anti-inflammatory and neuroprotective effects of omega-3s could contribute to the observed reductions in self-harm and suicidal ideation.

Our findings are consistent with the broader body of research highlighting the mental health benefits of omega-3 FA. Multiple studies have shown that omega-3 supplementation can be used as a supportive treatment for depression (13–15,17). For example, the Framingham Heart Study found significant associations between higher omega-3 levels and lower risks of major depressive disorder (37). A separate meta-analysis of RCTs showed that Omega-3 supplementation can be an effective treatment for bipolar depression (35). Additionally, multiple studies have linked Omega-3 FA to both suicide risk and risk of self-harm (18–23). Our study extends this knowledge by specifically focusing on self-harm and suicidal ideation, providing further evidence of the mental health benefits of omega-3s.

Given the associations observed in this study, increasing omega-3 intake through dietary sources or supplementation could be a valuable strategy in public health interventions aimed at reducing self-harm and suicidal behaviors. Public health policies promoting the consumption of omega-3-rich foods, such as fish and certain plant oils, may help mitigate the risks associated with self-harm and improve overall mental health.

Despite the robust findings, this study has several limitations. The observational nature of the study precludes causal inferences, however, we adjusted for numerous covariates of importance, and saw similar patterns in retrospective, cross-sectional and prospective analyses.

Additionally, self-reported data on self-harm and suicidal ideation may be subject to recall bias; even so, the strongest associations we identified were with medical record hard outcomes. The extent to which these findings can be generalized to other cultures or age groups is unclear.

Future research should aim to establish causality through randomized controlled trials and explore the optimal dosage and duration of omega-3 supplementation for mental health benefits.

In conclusion, this study provides evidence that higher levels of omega-3 FA, particularly DHA, are associated with reduced risks of self-harm and suicidal ideation. These findings highlight the potential of omega-3 FA as part of nutritional strategies to improve mental health and reduce self-harm behaviors. Further research is warranted to confirm these associations and explore their practical applications in mental health interventions.

## Data Availability

All data produced in the present study are available upon reasonable request to the authors but individual level data from participants in the UK Biobank can only be obtained via permission from the UK Biobank.

## Abbreviations

BMI: Body Mass Index
CI: Confidence Interval
CV: Cardiovascular
CVD: Cardiovascular Disease
DHA: Docosahexaenoic Acid
EPA: Eicosapentaenoic Acid
FA: Fatty Acid
FOS: Fish Oil Supplement
HR: Hazard Ratio
MDD: Major Depressive Disorder
NMR: Nuclear Magnetic Resonance
NSSI: Non-suicidal Self-injury
SD: Standard Deviation
UKBB: United Kingdom Biobank

## Declaration of Generative AI and AI-assisted technologies in the writing process

No AI or AI-assisted technologies were used in this writing process.

## Contributions of each author

WGF and ELO– assisted in writing, gathering information, and figures

NT and JW – assisted in informatics, tables, and writing of manuscript

JHO and WSH – concept, writing of manuscript, and finalization of manuscript

