## Supplementary Materials for "The Association Between Plasma Omega-3 Fatty Acids and Suicidal Ideation/Self-harm in the United Kingdom Biobank"

**Supplemental Table 1a – Variable definitions and UKBB questions/ICD-10 codes**

| **Response** | **UK biobank ID** | **Question/ICD9/ICD10** |
| --- | --- | --- |
| Life is not worth living | 20479 | Question asked: "Many people have thoughts that life is not worth living. Have you felt that way?" |
| Contemplate Self-harm | 20485 | Question asked: "Have you contemplated harming yourself (for example by cutting, biting, hitting yourself or taking an overdose)?" |
| Contemplate Self-harm 1yr | 20486 | Question asked: "Have you felt this way in the last 12 months?"  Question was asked when Field 20485 was any Yes. |
| Ever Self-harm (Self-Report) | 20480 | Question asked: "Have you deliberately harmed yourself, whether or not you meant to end your life? |
| 12 month Self-harm | 20481 | Question asked: "Have you harmed yourself in the last 12 months, whether or not you meant to end your life?"  Question was asked when Field 20480 was Yes. |
| Ever Self-harm (medical Record) | 41270  41271 | All ICD10 codes that start with X6,X7,X8  All ICD9 codes that start with E95 |
| First time Self-harm | 41270  41271  41280  41281 | All ICD10 codes that start with X6,X7,X8  All ICD9 codes that start with E95  Removed all subjects with prevalent self-harm events.  Cox proportional hazard model |
| Any incident Self-harm | 41270  41271  41280  41281 | All ICD10 codes that start with X6,X7,X8  All ICD9 codes that start with E95  Adjusted for prevalent self-harm events.  Cox proportional hazard model |

**Supplemental Table 1b. Variable definitions and correspondence to UKBB variable IDs - covariates**

| **Variables** | **UKBB IDs** | **Coding of UKBB** | **Details** |
| --- | --- | --- | --- |
| Age | 21022 |  | years |
| Anxiety | 20002 | 1287,1288,1469,1615,1614,1616,1243 |  |
| Alcohol | 1558 | Missing: NA, -3  Daily: 1  3-4x per week:2  1-2x per week:3  Rarely:4,5,6 |  |
| BMI | 21001 |  | Kg/m2 |
| Comorbidities | 20001  20002 | Any 20001  20002: 1065,1072,1111,1075,1074,1138,1139,1142,1143,  1220,1222,1223,1276,1224,1225,1226,1322,1112,1113,  1154,1081,1082,1086,1192,1193,1194,1067,1087,1079,  1078,1207,1277,1264,1263,1265,1114,1262,1261,1156,  1155,1134,1157,1158,1136,1309,1331 | Categorical:  0,1,2,3+ |
| DHA | 23457 |  | Percent of total fatty acids (NMR) |
| Doctor Visit for nerves, anxiety, tension, depression | 2090 |  |  |
| Education | 6138 | College: 1,5,6  High School: 2,3,4  Less than High School: -7 | College  High School  Less than High School |
| Employment | 6142 | Employed: 1  Retired: 2  Unemployed:5  Disabled: 4  Other: 6,7,-7,-3 |  |
| Ethnicity | 21000 | White:1001,1002,1003,1  Black:4001,4002,4003,4  Asian: 3001,3002,3003,3004,5  Other:2,6,2001,2002,2003,2004 | White, Black, Asian, Other |
| Exercise | 874/894/914  864/884/904 | Walking/Week: 874*864  Moderate Act./Week: 894*884  Vigorous Act./Week: 914*904  Weekly MET-like Exercise = 3.3*walking/week + 4*mod.act./week + 8*vig.act./week | Quartiles of Weekly Met-like Exercise  (four categories) |
| Marital Status | 6141, 709,670 | Married: 6141 = 1  Unmarried: 6141 <>1  Unmarried: 6141 is NA and  709 =1 Or 670: 4,5 | Married  Unmarried  Missing |
| Omega3 | 23451 |  | Percent of total fatty acids (NMR) |
| Psychotropic Meds | 20003 | 1140855856, 1140855914, 1140855920, 1140855930, 1140855944, 1140855960, 1140855976, 1140856040, 1140856052, 1140856074, 1140856092, 1140856130, 1140861026, 1140861042, 1140861046, 1140861054, 1140861066, 1140861072, 1140861078, 1140861082, 1140861086, 1140861096, 1140861104, 1140861110, 1140861120, 1140861124, 1140861134, 1140861144, 1140861156, 1140861160, 1140861170, 1140861180, 1140861184, 1140861188, 1140861192, 1140861204, 1140861210, 1140861216, 1140861222, 1140861224, 1140861230, 1140861236, 1140861240, 1140861250, 1140861260, 1140861270, 1140861280, 1140861290, 1140861300, 1140861310, 1140861320, 1140861330, 1140867184, 1140867208, 1140867218, 1140867244, 1140867272, 1140867288, 1140867304, 1140867306, 1140867312, 1140867342, 1140867398, 1140867406, 1140867414, 1140867420, 1140867444, 1140867952, 1140868120, 1140867632, 1140867658, 1140867668, 1140867690, 1140867712, 1140867818, 1140867824, 1140867852, 1140867858, 1140867876, 1140867884, 1140867888, 1140867490, 1140867494, 1140867498, 1140867504, 1140867518, 1140872150, 1140872228, 1140871986, 1140872228, 1140872290, 1140872172, 1140872186, 1140872304, 1140883494, 1140883482, 1140883518, 1140883588, 1140883656, 1140883658, 1140883662, 1140883670, 1140883858, 1140909722, 1140909798, 1140909806, 1140909800, 1140909802, 1140909804, 1140909816, 1140909818, 1140910704, 1140910374, 1140917132, 1140917138, 1140927460, 1140927956, 1140928946, 1140922638, 1140922638, 1140922638, 1141151978, 1141152732, 1141152848, 1141152860, 1141152736, 1141157336, 1141153490, 1141164244, 1141164246, 1141164248, 1141167976, 1141167932, 1141167940, 1141167026, 1141168396, 1141168398, 1141168436, 1141168444, 1141169714, 1141169722, 1141171932, 1141171940, 1141171578, 1141171482, 1141174500, 1141174508, 1141176854, 1141176858, 1141181616, 1141181708, 1141182732, 1141182592, 1141181508, 1141189134, 1141189868, 1141190158, 1141195974, 1141199446, 1141200564, 1141200004, 1141201792, 2018602634 |  |
| Sex | 31 |  | Female  Male |
| Smoking Pack Years | 20161  20116 | Pack Years: 20161  Smoking Status: 20116  Never:0  Previous:1  Current:2 | If Pack Years missing imputed to the mean for the subjects smoking status (0, 21.7, 27.6). |
| Townsend Deprivation Index | 22189 |  |  |

**Supplemental Table 2a. Self -harm – History of self-reported and medical record inferred**

|  |  |  | **History of Self Harm**  **(Self-Report) (N=80745)** | | **History of Self-Harm**  **(Medical Record) (N=258102)** | | |
| --- | --- | --- | --- | --- | --- | --- | --- |
|  |  | **n** | **% (x)** | **aOR (95% CI)** | **n** | **% (x)** | **aOR (95% CI)** |
| **DHA** | per IQ_5_R | 80745 | 4.1%(3350) | 0.96(0.87,1.05) | 258012 | 0.5%(1284) | 0.71(0.6,0.83)*** |
|  | Q1 | 13390 | 5.1%(681) | NA | 51587 | 0.8%(431) | NA |
|  | Q2 | 14981 | 4.2%(632) | 0.84(0.75,0.95)** | 51611 | 0.6%(305) | 0.87(0.75,1.02) |
|  | Q3 | 16217 | 3.9%(625) | 0.79(0.7,0.9)*** | 51591 | 0.4%(224) | 0.74(0.63,0.88)*** |
|  | Q4 | 17628 | 4%(712) | 0.88(0.78,1)* | 51617 | 0.3%(174) | 0.67(0.56,0.81)*** |
|  | Q5 | 18529 | 3.8%(700) | 0.89(0.78,1)* | 51606 | 0.3%(150) | 0.67(0.55,0.83)*** |
| **Total**  **Omega-3** | per IQ_5_R | 80745 | 4.1%(3350) | 0.96(0.87,1.05) | 258012 | 0.5%(1284) | 0.72(0.61,0.84)*** |
|  | Q1 | 14405 | 5.4%(780) | NA | 51597 | 0.8%(407) | NA |
|  | Q2 | 15308 | 4.3%(658) | 0.86(0.77,0.97)** | 51605 | 0.6%(315) | 0.97(0.83,1.13) |
|  | Q3 | 16192 | 3.9%(633) | 0.84(0.75,0.94)** | 51605 | 0.4%(215) | 0.74(0.62,0.89)*** |
|  | Q4 | 17106 | 3.8%(644) | 0.86(0.76,0.96)** | 51600 | 0.4%(197) | 0.8(0.67,0.96)* |
|  | Q5 | 17734 | 3.6%(635) | 0.9(0.8,1.01) | 51605 | 0.3%(150) | 0.69(0.57,0.85)*** |
| **ALA+EPA**  **+DPA** | per IQ_5_R | 80745 | 4.1%(3350) | 0.96(0.87,1.05) | 258012 | 0.5%(1284) | 0.75(0.65,0.88)*** |
|  | Q1 | 15364 | 5.4%(833) | NA | 51601 | 0.7%(378) | NA |
|  | Q2 | 15774 | 4.2%(669) | 0.88(0.79,0.98)* | 51598 | 0.6%(307) | 1.02(0.87,1.19) |
|  | Q3 | 15980 | 3.8%(614) | 0.85(0.76,0.95)** | 51606 | 0.5%(236) | 0.83(0.7,0.99)* |
|  | Q4 | 16548 | 3.9%(644) | 0.92(0.82,1.03) | 51600 | 0.4%(192) | 0.74(0.62,0.89)** |
|  | Q5 | 17079 | 3.5%(590) | 0.87(0.78,0.98)* | 51607 | 0.3%(171) | 0.74(0.61,0.9)** |

**Supplemental Table 2b. Self -harm – Previous 12-months self-reported and medical record inferred**

|  |  | **12-month Self Harm (Self Report) (N=80,745)** | | | **12-month Self Harm (Medical Record Inferred) (N=258,012)** | | |
| --- | --- | --- | --- | --- | --- | --- | --- |
|  |  | **n** | **% (count)** | **aOR**  **(95% CI)** | **n** | **% (count)** | **aOR (95% CI)** |
| **DHA** | per IQ_5_R | 80745 | 0.4% (300) | 1(0.73,1.37) | 258012 | 0%(90) | 0.52(0.28,0.98)* |
|  | Q1 | 13390 | 0.5%(61) | NA | 51587 | 0.1%(31) | NA |
|  | Q2 | 14981 | 0.4%(58) | 0.97(0.67,1.42) | 51611 | 0%(25) | 0.99(0.57,1.72) |
|  | Q3 | 16217 | 0.4%(67) | 1.13(0.78,1.63) | 51591 | 0%(14/) | 0.64(0.33,1.25) |
|  | Q4 | 17628 | 0.4%(62) | 1.06(0.72,1.55) | 51617 | 0%(15/) | 0.82(0.42,1.59) |
|  | Q5 | 18529 | 0.3%(52) | 0.98(0.65,1.47) | 51606 | 0%(5/) | 0.32(0.12,0.87)* |
| **Total**  **Omega-3** | per IQ_5_R | 80745 | 0.4%(300) | 0.91(0.67,1.23) | 258012 | 0%(90) | 0.58(0.32,1.07) |
|  | Q1 | 14405 | 0.5%(69) | NA | 51597 | 0.1%(28) | NA |
|  | Q2 | 15308 | 0.5%(70) | 1.14(0.81,1.61) | 51605 | 0%(19) | 0.85(0.47,1.55) |
|  | Q3 | 16192 | 0.4%(59) | 1.01(0.7,1.45) | 51605 | 0%(20) | 1.02(0.56,1.86) |
|  | Q4 | 17106 | 0.3%(55) | 1.01(0.7,1.47) | 51600 | 0%(21) | 1.29(0.71,2.35) |
|  | Q5 | 17734 | 0.3%(47) | 0.99(0.67,1.47) | 51605 | 0%(2) | 0.14(0.03,0.62)** |
| **ALA+EPA**  **+DPA** | per IQ_5_R | 80745 | 0.4%(300) | 0.88(0.65,1.2) | 258012 | 0%(90) | 0.69(0.39,1.22) |
|  | Q1 | 15364 | 0.5%(80) | NA | 51601 | 0%(23) | NA |
|  | Q2 | 15774 | 0.4%(70) | 1.03(0.74,1.44) | 51598 | 0%(23) | 1.29(0.71,2.35) |
|  | Q3 | 15980 | 0.3%(49) | 0.80(0.55,1.16) | 51606 | 0%(19) | 1.14(0.61,2.15) |
|  | Q4 | 16548 | 0.4%(61) | 1.07(0.75,1.52) | 51600 | 0%(18) | 1.2(0.63,2.29) |
|  | Q5 | 17079 | 0.2%(40) | 0.78(0.52,1.17) | 51607 | 0%(7) | 0.53(0.22,1.29) |
